# Evaluation of the Hypertension Surveillance System at Pilot Hypertension Prevention and Control Health Facilities in Addis Ababa, Ethiopia, 2022

**DOI:** 10.1101/2025.01.15.24314484

**Authors:** Nigus Goshim

**Affiliations:** SPHMMC

**Keywords:** Hypertension, Surveillance system evaluation, Addis Ababa, Ethiopia

## Abstract

**Introduction:** Hypertension is a major cause of premature death worldwide. Evaluating a surveillance system promotes the best use of data collection resources and ensures that systems operate effectively. It allows us to determine whether a specific system is useful for a particular public health initiative and is achieving the goals of the public health program and the data collection objectives. Ethiopian hypertension control initiatives were recently launched (2019) and implemented in limited health facilities. Therefore, this evaluation is aimed at the hypertension surveillance system of Addis Ababa, Ethiopia.

**Method:** A descriptive cross-sectional study was conducted from December 21 to 31, 2022. Eight health care facilities were evaluated based on updated CDC guidelines for evaluating public health surveillance systems, and one key informant from each facility was interviewed using semi structured questionnaire. The data were analyzed descriptively. The data were analyzed with Excel 2021 and summarized by the frequency mean, percentage and rate.

**Results:** The system detected cases (8/8) and monitored the control rate, attendance rate and missed visits. All interviewed staff (8/8) stated that the case definition, diagnosis and treatment algorism of hypertension were easy. Regarding acceptability, all staff believed that stakeholders were engaged, and 7/8 of the facilities recorded and reported data in a timely manner, but there was no acknowledgment given to the staff (0/8). The stability and data quality were 95% and 72.9, respectively. The representativeness was 24% to 36% based on prevalence, and regarding sex, it ranged from 88% to 108%. predictive positive value (31.4%) and zero flexibility.

**Conclusion:** The system was useful for detecting morbidities and retaining patients on treatment. It has good simplicity, acceptability and stability but requires improvement in its flexibility, representativeness, data quality and positive predictive value. The system is better able to scale up to other health facilities.

## Introduction

Hypertension is a disease in which blood pressure in blood vessels is constantly elevated. High blood pressure is a serious condition and can increase the risk of heart, brain, kidney and other diseases (1). The African region of the World Health Organization (WHO) has the highest prevalence of hypertension (27%), while the region of the WHO Americas has the lowest prevalence of hypertension (18%). The number of adults with hypertension (HTN) increased from 594 million in 1975 to 1.13 billion in 2015, with the increase occurring primarily in low- and middle-income countries. This increase is primarily due to increased risk factors for hypertension (HTN) in these populations (2).

In Ethiopia, the prevalence of HTN ranges from 7% to 37% (3). According to THE Ethiopia Noncommunicable Disease (NCD) 2015 step survey, the prevalence of increased blood pressure, i.e., systolic blood pressure (SBP) ≥140 and/or diastolic blood pressure (DBP) ≥ 90 mmHg, was 15.8%. It was 16.3% in females and 15.5% in males. It varies between urban and rural areas. The urban prevalence of increased blood pressure (BP) was 19.7%, and the rural prevalence was 14.9% (4). In Addis Ababa, many studies have shown that nearly one-third of adults are hypertensive (5–7). The stakeholders involved in hypertension prevention control are Resolve to Save Life, the World Health Organization, the Federal Ministry of Health, the Health Bureau, zonal sub city health department and health care facilities.

Hypertension is the most common cause of premature death worldwide. More than one in four men and one in five women are affected by the disease. The burden of hypertension (HTN) is felt disproportionately in low- and middle-income countries, where two-thirds of cases occur, largely due to the increase in risk factors in these populations in recent decades (1). Hypertension prevalence is higher in older adults, adults with lower family income, those with lower education, those with diabetes, those with obesity, and those with a disability than in their counterparts (8). The costs of HTN increase year to year. This indicates a growth in costs from year to year and a future increasing burden on society (9). Cardiovascular disorders such as coronary heart disease (CHD), congestive heart failure (CHF), ischemic and hemorrhagic stroke, renal failure, and peripheral arterial disease (PAD) are all twice as likely to occur in people with hypertension (HTN). It is often associated with additional CVD risk factors, and the risk of CVDs increases with the total burden of risk factors (10).

Bringing down HTN reduces heart attacks, strokes, renal damage, and other health issues. Hypertension can be prevented by reducing salt intake (to less than 5 g daily), eating more fruit and vegetables, being physically active on a regular basis, avoiding the use of tobacco, reducing alcohol consumption, limiting the intake of foods high in saturated fats and eliminating/reducing trans fats in the diet (1). Strategic interventions can prevent premature death from cardiovascular disease among people aged 30-69 years. Improving HTN control from 15% to 50% would save more than 640,000 lives each year, and reducing salt intake by 30% would save more than 720,000 lives each year. Eliminating trans fats would save more than 250,000 lives each year (11).

To prevent the impact of HTN, the World Health Organization (WHO) initiated the HTN prevention and control project, which is an extension of the existing WHO Package of Essential Noncommunicable Disease (WHO-PEN) interventions and HEARTS strategies. With the approval of the Ethiopian Federal Ministry of Health (FMOH), the Resolve to Save Lives (RTSL) launched the Ethiopian Hypertension Control Initiative (EHCI) in 2019. Resolve to Save Lives (RTSL) supported the development and implementation of evidence-based hypertension control programs in Bangladesh, China, Ethiopia, India, Nigeria, the Philippines, Thailand, Turkey, Vietnam, and 21 Pan America Health organization (PAHO) member countries, which treat more than 6 million people (11). The project has been implemented in seven (7) regions or cities of Ethiopia: Addis Ababa city, Amhara region, Diredawa city, Sidama region, Oromia region, Somali region and Tigray region. It is currently (2022) being implemented in 62 primary health care facilities (12).

The evaluation of a surveillance system promotes the best use of data collection resources and ensures that systems operate effectively. Surveillance system evaluation allows us to define whether a specific system is useful for a particular public health initiative and is achieving the overarching goals of the public health program and the data collection objectives (13). Ethiopian HTN control initiatives were recently launched (2019) and implemented in limited health facilities (12). There is no published evaluation on the HTN surveillance system in Addis Ababa. Therefore, evaluating the HTN surveillance system is important for assessing the strength and achievement of the system which is used to scale up the system to other primary health care facilities in the country. Therefore, this evaluation aimed to evaluate the hypertension surveillance system on pilot HTN prevention and control program health facilities in Addis Ababa, Ethiopia, 2022.

### Purpose and operation of the hypertension surveillance system

The main focus of the program is the prevention and management of HTN at the primary health care level through the standardization of protocols, capacity building of staff and robust monitoring of outcomes. In addition, the project included community-based behavior change communication on the reduction of salt intake at pilot sites (12). The scope of HTN prevention and control projects is improving the standard, access and quality of HTN programs in primary health care in Ethiopia. Its aim is to increase the enrollment of HTN patients in treatment, increase the blood pressure control rate and maintain care. It also monitors trends in HTN prevention and control by using standardized informed policy decisions and evaluating interventions so that effective and efficient actions/policies can be identified and supported (12).

Trained workers enter the patient’s data on simple application at the point of service care. When the data are entered into the simple application, the authorized body can see immediately. Resolve to Save Lives (RTSL) program managers, coordinators and ministry of health noncommunicable disease program stakeholders can access and monitor the data. The outpatient department directors or team leaders of each health care facility are able to access the data and monitor it. In addition, the health facilities report monthly via the District Health Information System (DHIS2).

### System components

The population under surveillance was all adults aged 30 years and older. The project is only run-on selected health care facilities. Healthcare workers (NCD staff) record every new patient’s sex, age, blood pressure (BP), comorbidities and drugs on simple application. Each patient has identification (ID) cards with unique QR codes. At each visit, patient information such as BP, drugs if changed and comorbidities from the available list can be updated. The simple application automatically sent personalized SMS/WhatsApp reminders to patients who reminded them to return for visits (14). The application shows lists of overdue patients with risk analysis. Risk analysis helps healthcare workers prioritize calling “high risk” patients first.

The NCD staff call the overdue patients to return to the health facilities. Managers receive daily reports to monitor progress, and patients can chart their own blood pressures and blood sugars (14). In addition to simple application, the NCD staff records patients’ information on the outpatient register, longitudinal register and defaulter tracing register. Noncommunicable disease staff collect monthly reports manually from the registries and submit them to the health management information system (HMIS) case team. The HMIS case team entered the data from the district health information system (DHIS2).

### Resources used to operate the system

The system needs trained health workers, drugs, medical equipment, laboratory services, internet connections, android phones, etc., Resolve to save life support (RTSL) support countries to choose a simple, proven treatment protocol, implement community-based care and task sharing, ensure a regular supply of medications, and support patient centered services that reduce barriers to adherence and use information systems to improve patient care.

## Methods

### Study area

The evaluation was conducted at the Addis Ababa RTSL sites, which are located in Gulele sub city and it is one of the 11 sub cities of Addis Ababa. According to Central statistical agency (CSA) estimates, the sub city has 384,649 people. There are seven (7) health centers and one (1) specialized hospital Pilot sites for HTN prevention and control program. The seven health centers serve 287,970 people. Saint Peter’s Specialized Hospital has no defined population. Females constituted 52% of the population.

This evaluation was conducted from December 21 to 31, 2022. The source populations were health facilities, hypertensive patients on follow-up and NCD OPD staff. The study population consisted of eight RTSL healthcare facilities, including seven health centers and one specialized hospital. Additionally, noncommunicable disease outpatient department (OPD) staff working within the RTSL sites and 80 selected records of hypertensive patients who had been taking antihypertensive drugs for at least three months were also included in the study population. There are eight healthcare facility RTSL sites in Addis Ababa. Since the number of healthcare facilities are small, all were evaluated. From each facility, one key informant was interviewed. To assess the adherence to the HTN treatment protocol from each facility, 10 patients, i.e., a total of 80 patients who were registered for HTN treatment and who stayed on treatment for at least three months, were selected randomly from simple application and audited.

### Definition of term

**Suspected HTN** was defined as individual presenting resting blood pressure measurements (based on the average of three readings) at or above 140 mm Hg for systolic blood pressure or greater than or equal to 90 mm Hg for diastolic blood pressure (15).

**Confirmed HTN** was defined as at least two occasions of resting blood pressure measurement (based on the average three readings) at or above 140 mm Hg for systolic blood pressure or greater than or equal to 90 mm Hg for diastolic blood pressure (15).

**Usefulness**: the contribution to the prevention and control of adverse health events, including an improved understanding of the public health implications of such events (13). In this evaluation, morbidity and mortality were assessed.

**The simplicity** of a public health surveillance system refers to both its structure and ease of operation (13). It was evaluated based on 12 criteria, including the ease of diagnosis and treatment algorithms, the simplicity of the HTN definition for healthcare professionals at all levels, the number of organizations that received the report, the duration of data collection, the list of comorbidities entered into the simple app, the number of risk factors recorded in the app and the cohort registry, and the reporting source.

**Flexibility** is the adaptability of surveillance systems for changing information needs or operating conditions with little additional cost in terms of time, personnel, or allocated funds (13). It was assessed by whether the reporting formats (simple app form and registers) could be used for other newly occurring health events (disease or comorbidity) without much difficulty or not, difficulty implementing any change in the existing procedure of case detection, reporting, and formats and interoperability with other systems such as DHIS2. It was assessed by three criteria/questions.

**Data quality** was assessed by evaluating the clearness and ease of filling out data collection formats for all the reporting sites, by checking missing and incomplete data collection and reporting forms, outliers in the data recorded, data validity issues, training status of reporting sites and supervision regularity. It was assessed by six criteria/questions.

**Acceptability** reflects the willingness of individuals and organizations to participate in the surveillance system (13). It was assessed by stakeholder (NCD foals, health professionals) acceptance and engagement in the surveillance activities, patient attendance rate, whether staff measured and recorded blood pressure for all patients aged 30+ presenting at the clinic or not, staff adherence to the standard protocol for treating patients diagnosed with HTN, and staff in the patient forms, register, and reporting form in a timely manner. It was assessed by seven criteria/questions.

**The sensitivity** of a surveillance system can be considered on two levels. First, at the level of case reporting, the proportion of cases of a disease or health condition detected by the surveillance system can be evaluated (13).

**Predictive value positive (PVP)** is the proportion of persons identified as having cases who actually do have the condition under surveillance (13). It was assessed by dividing the number of patients who started antihypertensive medication by the number of patients with raised BP.

**Representative**- the accuracy of describing the occurrence of a health-related event over time and its distribution in the population by place and person (13). It was assessed by the health service coverage of the site, the health-seeking behavior of the populations to the diseases under surveillance and by comparing the demographic distribution of surveillance data of patients seen at the clinic with other community-level studies. It was assessed by four criteria/questions.

**Timeliness** reflects the speed or delay between steps in a surveillance system (13). **Stability** refers to the reliability (i.e., the ability to collect, manage, and provide data properly without failure) and availability (i.e., the ability to be operational when needed) of the public health surveillance system (13). This was assessed by the following: simple application work properly without failure or not, system data storage device error causing the loss of data or not, lack of resources that interrupt the surveillance system due to inadequate staffing, and instances of data loss due to system breakdown. It was assessed by eight criteria/questions.

**The simple application** is an easy-to-use mobile application for healthcare workers to record BPs, blood sugars, and medicines at every patient visit in approximately 13 seconds. It has a web-based dashboard for officials and health system managers to monitor HTN control across facilities and regions (14).

#### Operational definition

Regarding the attribute assessment, a positive response and/or records above 75% of the criteria were rated as good. Positive responses and/or records between 50% to74% were rated as satisfactory, and those below 50% were rated as poor. Positive responses and/or records of 6/8, 4/8 and less than 4/8 were rated as good, satisfactory and poor, respectively.

The data were collected by a principal investigator by using a semi-structured questionnaire through document review and interviews with key informants, i.e., health facility NCD staff. The attributes assessed included usefulness, simplicity, data quality, flexibility, stability, acceptability, representativeness and positive predictive value according to the CDC updated guidelines for evaluating public health surveillance systems (13). The data collected by interviews at every level were crosschecked with available documents, and the completeness of the information after every interview was checked. The data were analyzed descriptively and are presented in text, tables and graphs. Continuous data were analyzed with Excel 2016 and summarized by the frequency mean, percentage and rate. Categorical qualitative data are summarized as frequencies.

The evaluation findings were submitted in both soft and print formats to Saint Paul’s Hospital Millennium Medical College (SPHMMC) School of Public Health, Department of Epidemiology, as well as to the Centers for Disease Control and Prevention (CDC) in soft copy. It was presented at SPHMMC.

## Results

### Supportive function of the HTN surveillance system

#### Manual and Protocol

All visited health facilities had either a hardcopy or a softcopy HTN treatment protocol. None of the 8 health facilities had a standard case definition for HTN. All evaluated sites except Saint Peter’s Specialized Hospital (SPSH) had a BP measuring guide that was posted on the wall.

#### Supervision and training

All eight health facilities were visited at least twice by both the sub-city health department and the RTSL staff. The RTSL coordinator calls the NCD staff when the number of overdue patients increases. Only three of the eight health facility health workers trained on the HTN treatment protocol. The training was provided by the RTSL. All health workers were oriented on simple application.

#### Resources

All health facilities (8) had electronic BP devices to measure blood pressure. Each health facility has one tablet. Resolve to Save Life recharge 300-birr airtime for each facility monthly. Only Saint Peter’s Specialized hospital (SPSH) has a desktop device in the noncommunicable disease clinic. All health facilities except Saint Peter’s specialized hospital had information education and communication materials such as posters.

### Core function of the HTN surveillance system

#### Availability of tests for case confirmation (secondary hypertension)

Urine analysis and fasting plasma glucose test were available in all 8 health facilities and hemoglobin or hematocrit test available at six of the facilities during the evaluation. Lipid profile, serum potassium and creatinine test were available in four, two and five health facilities respectively. Electrocardiography (ECG*) was available at Saint Peter’s Specialized Hospital (SPSH) and ECG is not standard services in health center level.

#### Data reporting and analysis

In all facilities, there was no shortage of reporting formats. In 3/8 health facilities, the report did not agree with the register. Report completeness was 100% in all health centers, but timeliness was difficult to assess. Simple application system show disaggregates data by sex and time. It also shows BP control status, missed visits, BP recording status and drug stocks. The health care workers can follow the progress in simple application. All health facilities did not show the data by place. Only one health center conducted trend analysis. In all the health facilities, the health information management system (HMIS) focal was responsible for the data analysis. There was no well-documented analysis in any of the health facilities, and they did not have or know the appropriate denominators.

#### Case detection, registration and medication

In all visited health facilities, there was a functioning diabetes mellitus (DM) and HTN treatment register. The treatment algorithm was available and displayed on the wall/desk in all health facilities except the SPSH. Blood pressure was recorded at every visit. Among audited record of hypertensive patients (80 patients), 70 (87%) started antihypertensive drugs for BP 140/90 and above. The majority (58, 72.5%) of patients started initial antihypertensive drugs (amlodipine) as recommended by the Ethiopian HTN treatment protocol, and the remaining 22 (27.5%) did not agree with the protocol. Fifty-six (70%) patients who were received initial doses agreed with the protocol, whereas the remaining 24 (30%) did not agree with the protocol.

Seven of the eight health facilities had designed Blood pressure (BP) corner, with 6 (six) of these functional during the evaluation. Staff members were assigned for patient/client counseling and BP measurement. Counseling materials were available, and the counseling system in all health facilities was provided individually. In all facilities, patient reminders and follow-up were functional. The simple application shows that overdue patients and health care workers call patients from the lists. There was also a system for tracking initial defaulters. The defaulters are traced by communicating with health extension workers based on addresses from family folders. ***Drug inventory system***

In each month, the amount of available drugs was filled in simple application, except SPSH. There was a shortage of hydrochlorothiazide (HCT) and lisinopril in one health center, i.e., the Mychew Health Centre in the previous quarter (1^st^ quarter of 2022), and of enalapril in one health center, i.e., Shegole, during the evaluation. There was not enough buffer stock for the core drugs for three months in all the health facilities. Lisinopril was stock out in all health centers during the evaluation. Drugs lists did not fill in simple application in SPSH and the status was not known.

### Performance of the Surveillance System

#### Usefulness of the HTN Surveillance System

All adults aged 30 years and older were eligible for the HTN surveillance system. In all eight facilities, the system detected the morbidity of HTN. In 2022, 1,535 new cases of HTN were recorded on the simple application in 8 (eight) facilities. The incidence of HTN in seven health centers was 1.09% in 2022. Saint Peter’s specialized hospital has no defined population, so the incidence or prevalence was difficult to assess. A total of 4,759 hypertensive patients were recorded since the program initiated, i.e., 2020. Approximately one-third (1451) of the cases were from the SPSH. Therefore, the overall usefulness in detecting cases is 100%. In terms of mortality, 2/8 of the facilities detected deaths. There were 8 (eight) deaths reported: seven from Entotofana and one from Mychew Health Centers. The two health centers recorded cases of death during defaulter tracing. Therefore, the system in detecting mortality was 33.3%.

Risk factors are available in the cohort register, so we can assess factors associated with HTN. In simple applications, we can monitor trend cases, BP control status, missed visit, overdue patients and drug stock. From the simple application dashboard, the average BP control status in seven health centers in the last six months was 56.4% and the attendance rate was 79.3%.

#### Simplicity

All (8) NCD-OPD staff believed that it is not difficult to use the diagnosis algorithm at all health professional levels, which is 100% simple for the diagnosis algorithm. Regarding the case definition of HTN, all (8) interviewed staff members said that it is easy for all-level health professionals to detect cases. All interviewed staff members believed that the treatment algorithm for HTN is easy for all levels of health professionals. All (8/8) respondents believed that additional data collected on a case is time consuming. On average, 10 to 15 minutes were required to complete all formats for each new patient. However, it takes less than two minutes to complete one new patient dataset on simple application. Program stakeholders can see and access information and aggregate reports from simple applications at any time. Therefore, there is no need to prepare reports to send to program managers and stakeholders. Only one organization, i.e., the higher level, needs the report. The NCD outpatient department submitted monthly and quarterly reports to the health information management system (HMIS) case team. The health information management system (HMIS) case team entered the reports via DHIS2. The health workers expected to fill patients’ data on simple application, outpatient and longitudinal/cohort registers. Overdue patients were registered on the defaulter tracing register. On average, 10 minutes were required to collect monthly reports. Only four comorbidities are available for simple application, i.e., DM, heart attack, stroke and kidney disease. No risk factors are available on the application. However, on the longitudinal register, six categories of risk factors are available. Cohort, outpatient and BP corner screening registers are the source reports. From 80 expected points, 72 were positive responses or records that support simplicity. Therefore, the overall simplicity was 90% (Table 1).

**Table 1:** Simplicity of the hypertension surveillance System in Addis Ababa, Ethiopia 2022.

| Criteria |  | Total/Average | Total positive response/records |
| --- | --- | --- | --- |
| Diagnosis algorithm is easy all level health professionals | Yes | 8 | 8 |
|  | No | 0 |  |
| Case definition of HTN is easy for case detection by all level health professionals | Yes | 8 | 8 |
|  | No | 0 |  |
| Treatment algorithm of HTN is easy for all level health professionals | Yes | 8 | 8 |
|  | No | 0 |  |
| Additional data collected on a case are time consuming | Yes | 8 | 0 |
|  | No | 0 |  |
| Average time to fill the formats |  | 15 minutes |  |
| Average time it takes to fill the new patient data on simple application |  | 90 seconds | 8 |
| Average time spent on collecting data for report |  | 11 minutes | 8 |
| Organizations which need to receive reports |  | 1 | 8 |
| List of comorbidities fill on the simple app- |  | 4 | 8 |
| Number of risk factor filled in the simple application |  | 0 | 8 |
| Number of risk factor filled in the cohort registers |  | 6 | 8 |
| Number reporting source |  | 3 | 8 |
| Total positive response or records |  |  | 72/80 (90%) |

#### Flexibility

On simple application, we can record drugs, BP records, and three comorbidities but cannot be used for newly occurring health events (disease or comorbidity) other than stroke, heart attack, kidney diseases and DM. All health workers believe that any change in the existing procedure of case detection, reporting, and formats is difficult to implement. Neither paper nor web-based surveillance forms are flexible enough to change if additional information is needed. Specifically, simple application and DHIS2 cannot be manipulated by health workers at the facility level. The simple application was not interoperable with DHIS2 until this evaluation was conducted. None of the 3 criteria were fulfilled, and the flexibility was zero.

#### Data Quality

All NCD staff believe that the data collection formats for HTN are clear and easy for all the data collectors to fill. There were no missing or incomplete data collection or reporting forms in the five facilities. The reporting formats were not easy or clear for those who did not have an orientation on HTN indicators and data elements. Raised BP and enrolled care interchangeable used in some health facilities. Some missing values and incomplete registers were observed in 3/8 of the health facilities. Outliers were observed in 1/8 of the facilities. In Saint Peter’s specialized hospital, Outlier’s data were collected. With respect to simple application, 63%, 63% and 60% of missed visits occurred from August to September 2022, respectively. There were sites where follow-up visits were reported as new cases. Data validity issues such as inconsistency of reports when checked from different sources were observed in 3/8 of the evaluated health facilities. In all the health facilities, the NCD OPD staff were not trained or supervised regularly on data quality. Only 2/8 were trained on data quality. Of the 48 responses, 35 were positive. The overall data quality was 72.9% (Table 2).

**Table 2:** Data quality of hypertension surveillance system in Addis Ababa, Ethiopia, 2022.

| Criteria |  | Total | Total positive response/records | Status |
| --- | --- | --- | --- | --- |
| Format clearness and easiness | Yes | 8 | 8 | Good |
|  | No | 0 |  |  |
| Incomplete and missing of reporting form | Yes | 3 | 5 | Satisfactory |
|  | No | 5 |  |  |
| Presence of outliers in data recorded | Yes | 1 | 7 | Good |
|  | No | 7 |  |  |
| Presence of data validity issues | Yes | 3 | 5 | Satisfactory |
|  | No | 5 |  |  |
| Training for NCD | Yes | 2 | 2 | Poor |
|  | No | 6 |  |  |
| Percent of reports which are complete | Percent | 100 | 8 | Good |
| Total positive response/record |  |  | 35/48(72.9%) | Satisfactory |

#### Acceptability

All (8) interviewed NCD OPD staff believed that all stakeholders accepted and were well engaged in the surveillance activities. The stakeholders were willing to use the system in an especially comfortable manner with simple application. Newly assigned healthcare workers who worked in NCD-OPDs were not well engaged in HTN surveillance activities for many reasons. Some of the reasons were a lack of understanding of the relevance of the data to be collected, information gaps and report formats are time consuming. In 6/8 of the facilities, blood pressure was measured and recorded for all patients aged 30+ who presented at the clinic. All NCD-OPD staff members adhered to the standard protocol for treating patients diagnosed with HTN. Staff members fill out patient forms, registries, and reporting forms in a timely manner most of the time in 7/8 of the facilities. There was no special acknowledgment given to the NCD OPD staff in any of the facilities. The average 6-month attendance rate and control rate in the 7 health centers were 79.3%. Among 42 responses/or records, 37 supported acceptability, and the overall acceptability was 88% (Table 3).

**Table 3:** Acceptability of the hypertension surveillance in Addis Ababa, Ethiopia, 2022.

| Criteria |  | Total | Positive response/record |
| --- | --- | --- | --- |
| All the stakeholders accept and well engaged to the surveillance activities | Yes | 8 | 8 |
|  | No | 0 |  |
| Blood pressure measured and recorded for all patients aged 30+ presenting at the OPD | Yes | 6 | 6 |
|  | No | 2 |  |
| Staff fill in the patient forms, register, and reporting form in a timely manner | Yes | 7 | 7 |
|  | No | 1 |  |
| Acknowledgment by the system managers of the contributions made by the people who provided the data | Yes | 0 |  |
|  | No | 8 | 0 |
| Staff adhere to the standard protocol for treating patients diagnosed with HTN | Yes | 8 | 8 |
|  | No | 0 |  |
| Patients' attendance rate in the last 6 months |  | 79% | 8 |
| Total positive response/record |  |  | 37/42(88%) |

#### Representativeness

The health services coverage of the sub-city is 100%. All interviewed staff members believed that the populations under surveillance, i.e., all adults older than 30 years, had good health-seeking behavior for HTN. All (8) respondents said that health facility visitors are well represented by the system. Total positive response/record was 30.6/40. (Table 4). Among the new patients, 928 (60.46%) were female. Among the 85,875 individuals screened, 4,807 (5.6%) had raised BP, and 1,510 (31.4%) had HTN from raised BP. The representativeness in terms of raised BP was 36% (5.6%/15.6%) compared with the BP value reported in the national steps survey.

**Table 4:** Representativeness of the hypertension surveillance system in Addis Ababa, Ethiopia, 2022.

| Criteria |  | Total | Positive response/record |
| --- | --- | --- | --- |
| The health service coverage of the sub-city | Percent | 100 | 8 |
| The populations under surveillance have good health seeking behavior for HTN | Yes | 8 | 8 |
|  | No | 0 |  |
| Who do you think is well represented by the surveillance data? | health facilities visitors | 8 | 4 |
| Is the demographic distribution of patients seen at the clinic similar to that in the community? | Female | 60.46% | 8 |
|  | Male | 49.54% |  |
| Raised BP prevalence |  | 5.6% | 36% (2.6/8) |
| Total positive response/record |  |  | 30.6/40 (76.5%) |

#### Stability

The application demonstrated consistent functionality across all eight evaluated sites. No data loss occurred due to storage device errors. While minor issues, such as difficulties scanning BP passports, were encountered at one health center, overall performance was high. Staff turnover impacted service delivery at one facility, including Saint Peter’s Specialized Hospital. This resulted in a slight interruption of services. The total positive response and records was 39/40 (97.5%) (Table 5).

**Table 5:**
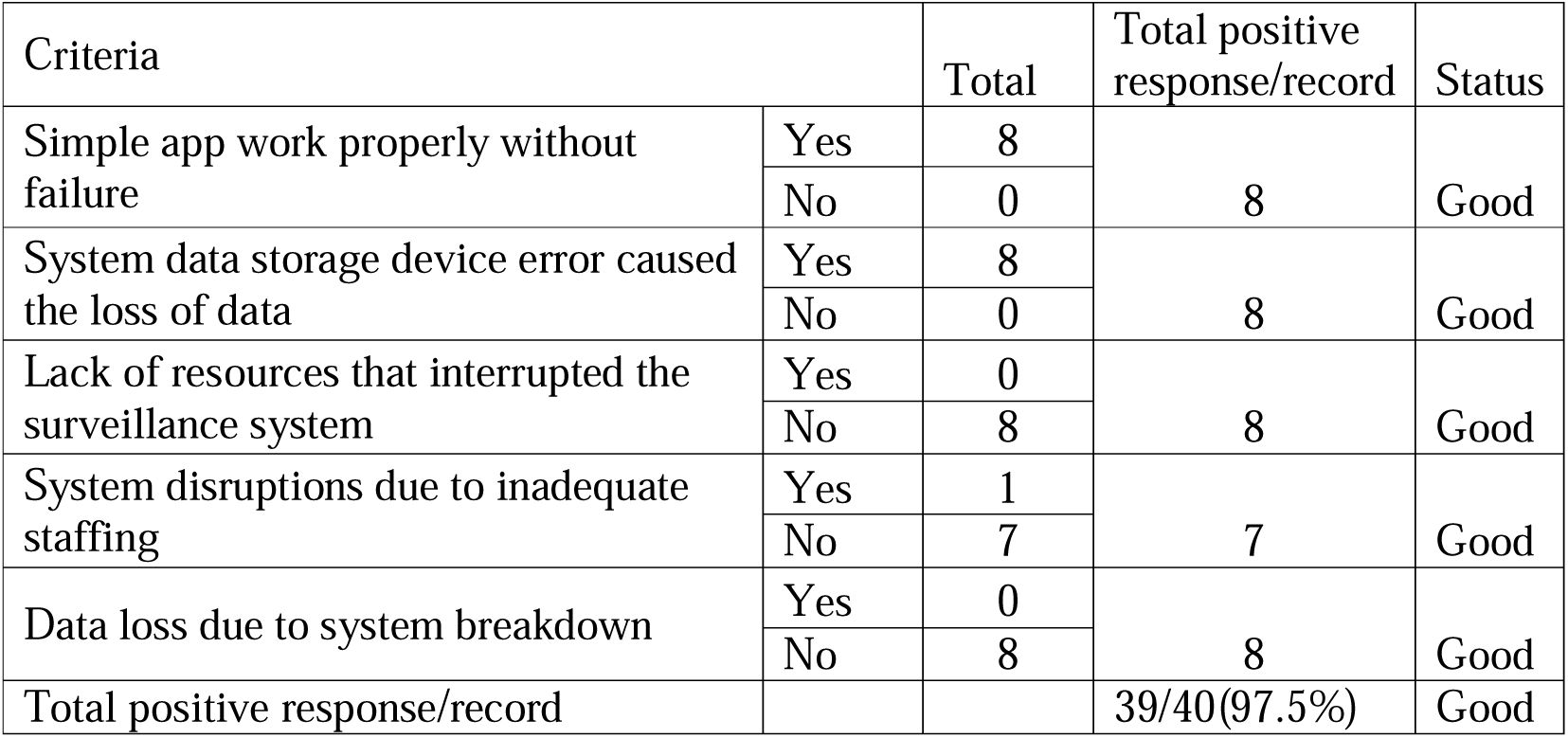
Stability of the hypertension surveillance system in Addis Ababa, Ethiopia, 2022.

| Criteria |  | Total | Total positive response/record | Status |
| --- | --- | --- | --- | --- |
| Simple app work properly without failure | Yes | 8 | 8 | Good |
|  | No | 0 |  |  |
| System data storage device error caused the loss of data | Yes | 8 | 8 | Good |
|  | No | 0 |  |  |
| Lack of resources that interrupted the surveillance system | Yes | 0 | 8 | Good |
|  | No | 8 |  |  |
| System disruptions due to inadequate staffing | Yes | 1 | 7 | Good |
|  | No | 7 |  |  |
| Data loss due to system breakdown | Yes | 0 | 8 | Good |
|  | No | 8 |  |  |
| Total positive response/record |  |  | 39/40(97.5%) | Good |

Regarding with simple application, it worked properly without failure in all evaluated sites (8/8). From 8 sites, only one health center faced a minor problem (failure to scan the BP passport). Storage device errors did not cause the loss of data (8/8). Trained health worker turnover caused an interruption in one facility (1/8). In Saint Peter’s specialized hospital, there was interruption of services due to staff turnover. Of the 40 responses, 39 were positive response or records and the stability was 97.5% (Table 5).

### Positive Predictive Value

Among the 85,875 screened adults, 4,807 (5.6%) were suspected to have HTN (raised BP). Among the patients with suspected HTN, 1,510 (31.41%) had confirmed HTN. True positives are confirmed HTN cases, and the sum of true positives and false positives is raised in BP cases. Therefore, the PPV calculated by the number of confirmed HTN cases divided by the number of raised BP cases multiplied by 100% From the available information, the predictive value of each facility is mentioned.

#### Timeliness

When health workers enter new patients’ data in a simple application, all stakeholders are able to see the data immediately. On simple application, 7/8 facilities entered patient data in a timely manner. The NCD OPD staff collected data from tally and different registers and submitted the HMIS case team monthly. There were no records or time logbooks for checking monthly and quarterly report timelines.

## Discussion

Eight health facilities were evaluated by using the CDC surveillance system evaluation framework. The evaluation showed that the system was useful for detecting morbidity but poor for detecting mortality. The system had good simplicity, satisfactory data quality, good acceptability and good stability but poor representativeness, low predictive positive value and no flexibility.

Antihypertensive drug treatment was started for BP <140/90 in 10(13%) of the cases. One reason was that some patients started antihypertensive drugs at other health facilities, controlled their BP and continued at the evaluated sites. Even if the Ethiopia HTN treatment protocol does not recommend antihypertensive drugs for individuals with a BP <140/90 mmHg, the WHO guidelines recommend antihypertensive treatment for individuals with cardiovascular disease with a systolic blood pressure of 130–139 mmHg (16). There were no fixed-dose combinations of antihypertensive drugs in any of the evaluated health facilities. Studies have shown that fixed-dose combinations have good adherence and better treatment outcomes (16–19). According to the Ethiopia HTN protocol, the first-line drug is amlodipine 5 mg (20). In this evaluation, 58 (72.5%) patients started with amlodipine, and 24 (30%) patients were prescribed initial doses that agreed with the protocol. This may be due to the use of different guidelines, such as standard treatment guidelines (STGs), Ethiopia primary health clinical guidelines (EPHCGs) and HTN treatment protocols. Each guideline has no clear instructions on dose or first-line drugs.

The system effectively identifies individuals who missed their scheduled appointments and categorizes them based on their risk level. This enables health workers to prioritize their outreach efforts to those at highest risk, ensuring timely interventions and improved adherence to treatment plans. Additionally, the system sends automated reminders to patients, further reducing the likelihood of missed appointments and defaulters. These features collectively contribute to enhanced adherence control rates and improved health outcomes. According to my evaluation, the control rate was 56.4%, which was better than that reported in other studies conducted in Jimma (42.8%) (21), North Ethiopia (51.4%) (22), Northwest Ethiopia (42.9%) (23), Southwest Ethiopia (50.3%) (24) and Shashemene, Ethiopia (40.3%) (25).

The diagnostic algorithm, case definition, and treatment algorithm were all easy to understand and use (8/8). Patient data were quickly and easily entered into the simple application (8/8). However, it was difficult to collect additional data to fill out different forms, such as OPD and longitudinal registers (0/8). A limited number of data elements are available for simple applications that are simple to fill. Overall, 72 out of 80 expected points were positive responses or records that supported the simplicity of the system, resulting in an overall simplicity rating of 90%. The case definition of this evaluation was rated as good (8/8), which was better than the evaluation conducted in Kenema Government Hospital, Sierra Leone, in 2021 (68.7%), which was assessed more sample (26).

The data quality of this evaluation was rated as satisfactory (72.9%) , which was slightly higher than that of the evaluations conducted in the Pulo Lor Primary Healthcare Centre, Indonesia (66.9%) (27), and Kenema Government Hospital, Sierra Leone, which was poor (4.5%) (26). Acceptability was rated as good interims of stakeholders’ engagement (8/8), BP was measured for all eligible patients and patients (6/8), adherence to the HTN protocol (8/8), and timely recording and reporting of data (7/8); however, the acknowledgment of the staff was poor. The overall acceptability was 88%. The acceptability of the recording and reporting data in a timely manner was good (7/8), which was lower than that of the evaluation conducted at Kenema Hospital (100%)(26). The overall stability of the system was good (97.5%). The system did not interrupt due to resources (8/8), which was better than that at Kenema Hospital (64.1%) (26).

The representativeness was rated as good based on the health services coverage (100%), health seeking behavior of the population (8/8) and representativeness of the surveillance data (8/8). Among the new patients, 928 (60.46%) were female, which was similar to that reported in a study conducted in the Addis Ababa public health facility (58.3%)(7). Therefore, the representativeness is 104% (60.46/58.3). A study conducted at Zewditu Hospital revealed that females were higher (55.6%) composition (28) which give the representativeness 108% (60.46/55.6).

Another study conducted at Yekatit 12 Hospital in Addis Ababa show males were higher (53.8%) composition (29). Therefore, the representativeness was 112% (60.46/53.8), or 88% for females. This may be due to sample variability. In the three studies, females were slightly more represented in two of the studies, and males were slightly more represented in one study. The representativeness of the surveillance system regarding sex was good, ranging from 88% to 108%.

The prevalence of raised BP in this evaluation was 5.6%, which was much lower than that reported in the Ethiopia Step Survey 2015. The prevalence of raised BP was 15.6% nationally and 22.6% in Addis Ababa (30). A systemic meta-analysis showed that the prevalence of HTN among the Ethiopian population was estimated to be 19.6%, and it was greater in the urban population, i.e., 23.7%. In this evaluation, the representativeness (for prevalence) was 36% (5.6/15.6) compared with the national figure (15.6%), whereas it was 24% (5.6/23.7) compared with the urban prevalence of raised BP. The system representativeness on prevalence is low. This was because patients have many alternative health facilities for visiting and diagnosing patients outside the evaluated sites. The representativeness of this evaluation based on prevalence (24% and 36%) was better than that of the evaluation conducted in Primary Health Care in Bogor City in 2018, Indonesia, which was 15.8% (31).

In this evaluation, 31.4% of patients with raised BP started antihypertensive drugs, whereas in 2015, 9% and 2.1% of patients in Addis Ababa and nationally, respectively, started antihypertensive drugs (30). Even if the predictive positive value (PPV) of the system is low, it is higher than that of the step survey results. This may be due to sample differences and population sources. In this evaluation, the source was the health facility, whereas in the step survey, the source was the community. The PPV was lower than that reported in an evaluation conducted in the Pulo Lor Primary Healthcare Centre, Indonesia, which was 59.4% (27).

The scope of the evaluation was limited because not all attributes were assessed. Timeliness could not be fully evaluated due to the absence of a time logbook for the report. Timeliness can be verified using DHIS2, but the system was recently updated. Some reports were created in the old DHIS2 version, while others were created in the new version. Consequently, assessing report timeliness was challenging. In addition, sensitivity was not assessed due to the lack of a gold standard. If an individual has no raised BP, there is no chance to record again. Only raised BP was measured repeatedly. The system helps to improve attendance and trace defaulters, which are good for adherence. The system was useful for detecting morbidities and retaining patients on treatment. It has good simplicity, acceptability and stability but requires improvement in its flexibility, representativeness, data quality and positive predictive value. The system is recommended to scale up to other health facilities.

## Data Availability

All data produced in the present work are contained in the manuscript.

## Acknowledgments

I also acknowledge the Ethiopia Public Health Institute, Addis Ababa Health Bureau, Addis Ababa Regional Public Health Institute and Gulele Health Department for providing supporting letters. I acknowledge all the health facilities for allowing me to conduct this evaluation and for their good approach.

I would like to express my gratitude to my academic mentor Aman Yesuf (MPH/E, Assistant Professor), program mentor Henok G/Yohannes (MPH) and other personnel who supported me in the system evaluation.

## Abbreviations and Acronyms

CDC: Center of Disease Control and Prevention
CHD: Coronary Heart Disease
CHF: Congestive Heart Failure
CVDs: Cardiovascular Diseases
DBP: Diastolic blood pressure
EPHI: Ethiopia Public Health Institute
EHCI: Ethiopian Hypertension Control Initiatives
FBG: Fasting blood glucose
HEARTS: Healthy Evidence Access Risk Tool Protocol
HMIS: Health Management Information System
HTN: Hypertension
NCD: Non-Communicable Disease
PAD: Peripheral arterial disease
PAHO: Pan America Health Organization
PEN: Package of Essential
RTSL: Resolve to Save Lives
SBP: Systolic blood pressure
U/A: Urine analysis
WHO: World Health Organization

## Availability of data and materials

Most data are available in the paper. Additional data will be shared upon request.

## Ethics approval and consent to participate

Prior to conducting the evaluation, a concept note was submitted to the Ethiopia Public Health Institute (EPHI) for review and approval. Official support letters were obtained from EPHI, the Addis Ababa health bureau, and the Gulele sub city health department. All participants were informed and consent was obtained.

## Authors’ contributions

Nigus Goshim designed the study and conducted research at the field levels, analyzed the data, and prepared the manuscript.

## Consent for publication

Not applicable.

## Author information

^1^St. Paul’s Hospital Millennium Medical College, School of Public Health, Department of Field Epidemiology, Addis Ababa, Ethiopia.

